# Corticosteroid Prescribing Practices at Dermatology Outpatient Departments of Tertiary Care Hospitals: A Multicenter, Cross Sectional Study

**DOI:** 10.1101/2022.06.19.22276595

**Authors:** Nagina Sultana, Jannatul Ferdoush, Wafa Sarwar, Sharif Mohammad Towfiq Hossain, Arifa Hossain, Mosammat Asma

## Abstract

**Background:** In dermatology, corticosteroids are the most frequently prescribed medication group. The goal of this study was to examine the prescription pattern of glucocorticoids in patients with skin diseases in outpatient dermatology departments (OPD) in tertiary care hospitals.

**Methods:** A prospective, multi-centered, cross-sectional study was undertaken in the dermatology department of government affiliated private medical College hospitals in Chittagong, Bangladesh, from March 2021 to April 2022. A total of 450 prescriptions from dermatology out-patient department patients with skin diseases were obtained and evaluated for demographics, skin diseases, and corticosteroid use patterns.

**Result:** Out of 450 patients, majority belonged to the age group 21-40 years (44%) and were female (56%). The most frequent indication for prescribing corticosteroids were eczema (20%) urticaria (17%) and allergic Contact dermatitis (7%) and psoriasis (7%). The commonest corticosteroids used were betamethasone (19%) and Clobetasol proprionate (17%), Deflazacort (15%). Use of topical steroid combinations with Fusidic acid and Miconazole nitrate were prevalent, (4%) and (5%), respectively. Highly potent steroids were used in 51% cases whereas only 21% were given low potency steroids.

**Conclusion:** Physicians prefer highly potent corticosteroids, according to this study. To improve prescribing, physicians should receive ongoing medical education and be made aware of the irrational use of steroids. Researchers and policymakers can use the baseline data acquired in these studies to enhance judicial prescribing practices. Keywords: Corticosteroids, dermatology, prescribing pattern.

## Introduction

With the advent of topical corticosteroids, the current era of dermatotherapy began, and since then, these agents have been used in a variety of ways, with varying degrees of success. Many people suffer from typical skin disorder that affect people of all ages. Corticosteroids has significant role in the treatment of a various disorders, including skin problems. In topical form, it is likely to have more applications in dermatological therapy. Consultants frequently prescribe these medicines due to their potent immunosuppressive and anti-inflammatory effects. Interestingly, the similar processes that cause their beneficial anti-inflammatory characteristics are also responsible for their negative effects [1].

Acute urticaria, pemphigus vulgaris, keloid, fixed drug eruptions, eczema, and other dermatological conditions have all been treated with them. As a result, dermatologists have risen to the top of the list of corticosteroid prescribers [2]. However, caution must be taken during the selection process as well as with regard to dose regimes. Corticosteroids are often used to treat psoriasis, as well as adult atopic dermatitis and eczema [3]. Despite their usefulness, their inappropriate and extended use in a variety of dermatological disorders puts a patient at risk for a variety of side effects, ranging from minor to severe [4].

Corticosteroids’ most serious side effect is immunosuppression, which might make you more susceptible to bacterial and fungal infections. The side effects of topically administered corticosteroids include adrenal suppression, rosacea induced by steroid, dermal and epidermal thinning, striae, candidiasis (perioral), and hypertrichosis [5]. As a result, doctors should prescribe the lightest corticosteroid feasible to treat dermatological disorders [6].

A correct diagnosis, as well as consideration of the routes of application, frequency of administration, duration of therapy, and adverse effects, are all necessary for successful treatment [7].

Because the medicine is administered directly to the target organ, topical treatments are preferred. The dose can be easily adjusted based on the reaction [8]. Glucocorticoids (topical and systemic) are widely used alone or in combination with other medications [9]. To acquire the best possible benefits at the minimum possible therapeutic dose for the short period at a reasonable cost and with fewer side effects, it is critical to employ the safest and smallest number of steroids feasible [10]. This will allow for more sensible medicine use. Prescribing medications is a crucial skill that must be examined and provide feedback to physician on a regular basis. Since it indicates a physician’s diagnostic ability and attitude toward picking the most appropriate and cost-effective treatment. Their sensible usage, on the other hand, can help to reduce the cutaneous and systemic negative effects that corticosteroids might cause [7]. Physicians should prescribe steroid that will treat the skin related problem with the most therapeutic benefit and the fewest side effects possible [11].

Because of the high disease prevalence and the significant economic burden of skin disease treatment, it is critical to investigate skin disease drug prescribing patterns. Taking these facts into account, current study was conducted to evaluate corticosteroid medication prescribed in dermatology to create baseline data and evaluate various elements of drug prescribing procedures.

## Materials and Methods

A prospective, multicenter, cross-sectional study was done in the Dermatology department of the government affiliated private medical college of Chittagong, Bangladesh from March, 2021 to April, 2022. Data were collected from outpatient department of dermatology of BGC Trust Medical College, Chattogram Maa-O-Shishu Medical College, and Marine City Medical College. The protocol has been approved by the BGC trust medical college’s institutional review board.

### Study procedure

A total of 450 prescriptions of patients with skin problems from the dermatology out-patient department who were on corticosteroids and were correctly selected by the dermatologist of the relevant medical college were obtained and analyzed. A sufficient number of data collectors were engaged to collect the data. The information was gathered prospectively through direct observation in a specially developed pro forma that included demographic, dermatological disease, and pharmacological information. The prescriptions were examined for the following information: Diagnosed skin diseases, medicine information (corticosteroid alone or in combination with other medicines combination, topical, systemic corticosteroids, and potency).

### Statistical Analysis

The obtained data was numerically coded and imported into Microsoft Excel 2010, where it was evaluated while keeping respondents’ anonymity and privacy in mind. Microsoft Excel was used for statistical analysis, and data was evaluated as frequency and percentages.

## Result

The study included 450 prescriptions with corticosteroid prescriptions. The majority of the patients (196/450, 44 %) were belongs to 21-40 years age group. Females comprised about 56% of the patients (252/450) (Table I)

**Table I:** The demographic profile of the participants

| Item | No of patients (n=450) |  |
| --- | --- | --- |
| Age | Frequency | Percentage |
| <20 | 88 | 19% |
| 21-40 | 196 | 44% |
| 41-60 | 105 | 23% |
| >60 | 61 | 14% |
| Gender |  |  |
| Male | 198 | 44% |
| Female | 252 | 56% |

Table II demonstrated that the most prevalent indications for corticosteroid prescription were eczema (20%), urticaria (17%), milia rubra (8%), allergic contact dermatitis (7%), and psoriasis (7%).

**Table II:** Dermatological conditions indicated for Corticosteroid

| Dermatological Indication | Total No of patient (N=450) |  |
| --- | --- | --- |
|  | Frequency | Percentage |
| Eczema | 86 | 19 |
| Urticaria | 77 | 17 |
| Psoriasis | 33 | 7 |
| Lichen planus | 12 | 3 |
| Allergic Contact dermatitis | 31 | 7 |
| Pemphigus vulgaris | 6 | 1 |
| Adverse drug reaction | 22 | 5 |
| Seborrheic dermatitis | 23 | 5 |
| Keloid | 7 | 2 |
| Alopecia | 31 | 7 |
| Discoid lupus erythematosus | 3 | 1 |
| Planter keratoderma | 10 | 2 |
| Atopic dermatitis | 24 | 5 |
| Plurigo nodularis | 11 | 2 |
| Plurigo simplex | 17 | 4 |
| Miliaria rubra | 26 | 6 |
| Melasma | 31 | 7 |

Table III demonstrated that the most commonly used corticosteroids were betamethasone (86/450, 19%), clobetasol proprionate (77/450, 17%), and deflazacort (67/450, 15%).

**Table III:** Corticosteroids commonly recommended in dermatological outpatient department.

| Corticosteroid | Total N=450 |  |
| --- | --- | --- |
|  | Frequency | Percentage |
| Betamethasone | 86 | 19 |
| Clobetasol propionate | 77 | 17 |
| Deflazacort | 67 | 15 |
| Prednisolone | 49 | 11 |
| Mometasone furoate | 33 | 7 |
| Hydrocortisone | 48 | 11 |
| Flucinolone acetonide | 38 | 8 |
| Triamisolone | 34 | 8 |
| Halobetasol | 18 | 4 |

Table IV demonstrated that (356/450) 85% corticosteroids were administered alone. The most often used topical steroid combination is with Miconazole Nitrate (23/450, 5%), Fusidic acid (20/450, 4%), Gentamycin (3/450, 1%), and Clotrimazole (21/450, 5%). Over half of the steroids administered (230/450) were very potent, with just (115/450, 26%) being low potent. In terms of duration of use, 67 % (301/450) of patients had used corticosteroids for more than two weeks. Corticosteroid routes of administration were predominantly topical (67%) and parenteral (7%;34/450).

**Table IV:**
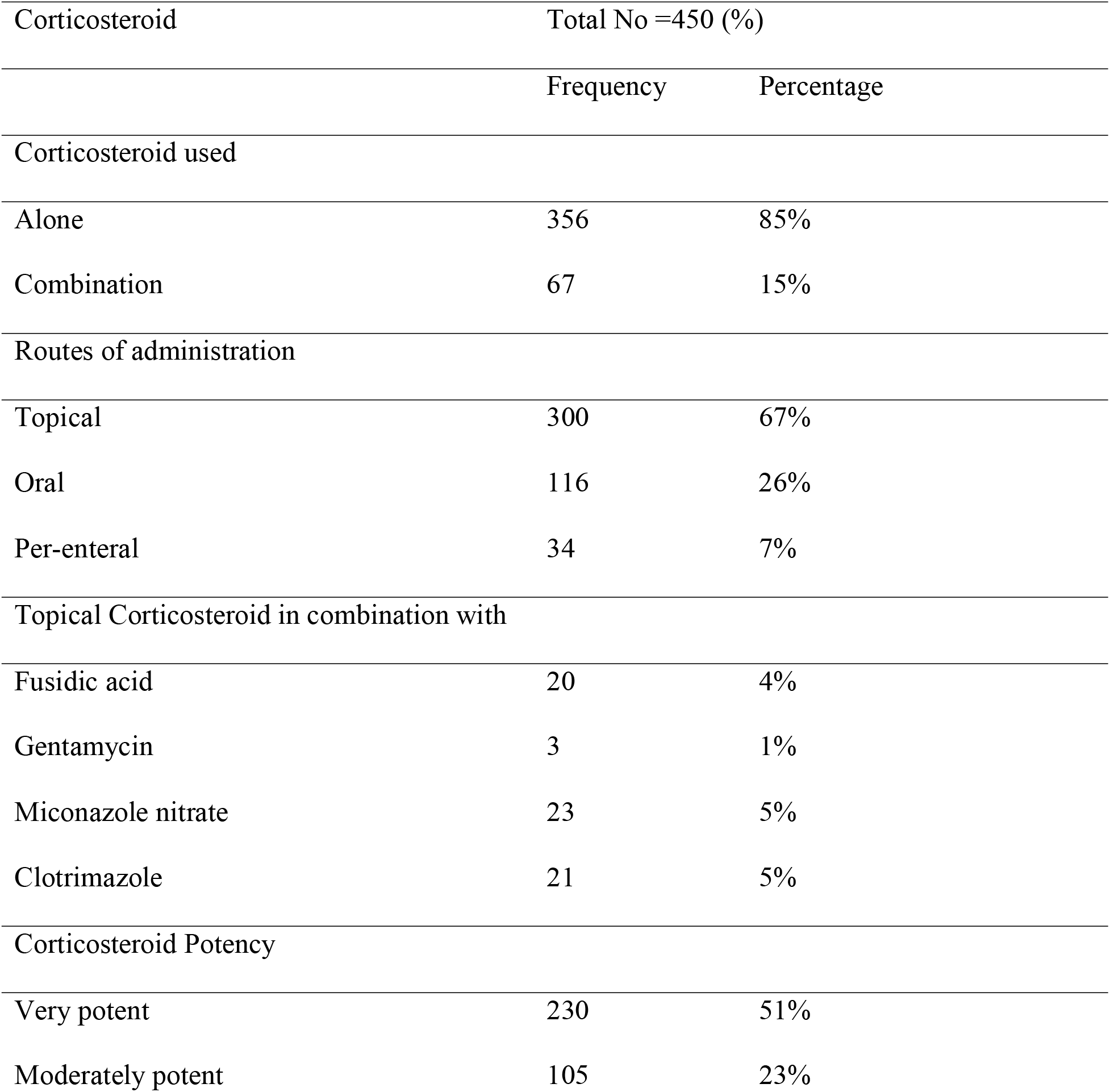
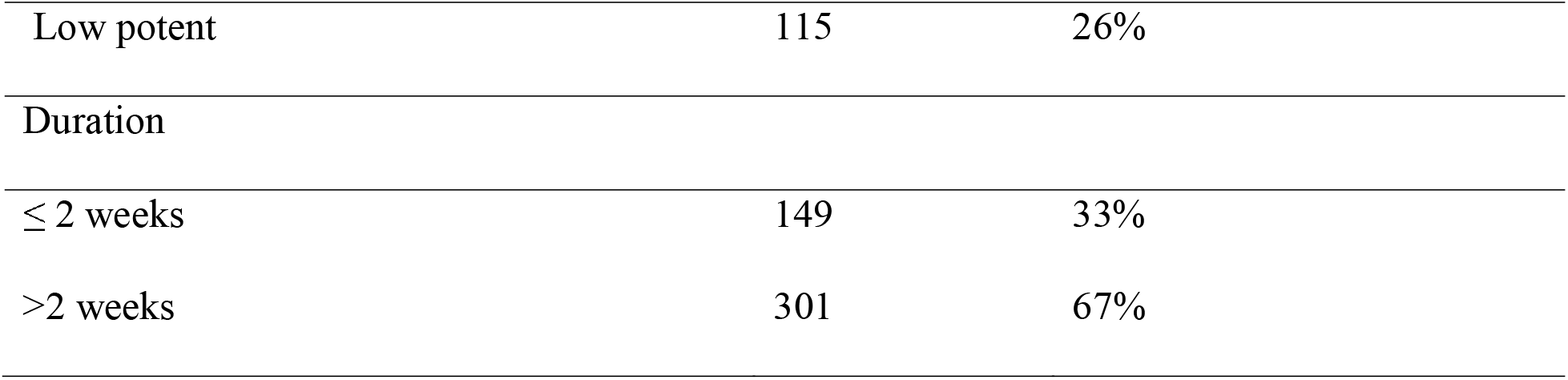
Detailed corticosteroid prescribed.

| Corticosteroid | Total No =450 (%) |  |
| --- | --- | --- |
|  | Frequency | Percentage |
| Corticosteroid used |  |  |
| Alone | 356 | 85% |
| Combination | 67 | 15% |
| Routes of administration |  |  |
| Topical | 300 | 67% |
| Oral | 116 | 26% |
| Per-enteral | 34 | 7% |
| Topical Corticosteroid in combination with |  |  |
| Fusidic acid | 20 | 4% |
| Gentamycin | 3 | 1% |
| Miconazole nitrate | 23 | 5% |
| Clotrimazole | 21 | 5% |
| Corticosteroid Potency |  |  |
| Very potent | 230 | 51% |
| Moderately potent | 105 | 23% |
| Low potent | 115 | 26% |
| Duration |  |  |
| $\leq 2$ weeks | 149 | 33% |
| $>2$ weeks | 301 | 67% |

## Discussion

Corticosteroids are among the widely used classes of medicine used in dermatological disease as both short-term and long-term treatments, although they are accompanied with a multitude of side effects [11]. Prescriptions should be audited on a regular basis to improve prescribing quality, decrease side effects and prescribing errors, provide criticism to physicians, and implementation of standard treatment guideline. The study goal was to gather baseline data by assessing corticosteroid prescribing patterns and evaluating monotherapy and the use of co-administered medicines in conjunction with corticosteroids in skin diseases.

The majority (44%) of the patient in this study were between the ages of 21 to 40, 56% were female. This is similar to the study conducted by Bylappa bk et al [12].

According to the current study, eczema (20%), urticaria (17%), miliaria rubra (8%), allergic contact dermatitis (7%), and psoriasis (7%) were the most common reasons for corticosteroid prescription.

Study conducted by Bylappa et al revealed contact dermatitis (47%) and psoriasis (14%) were the common dermatological condition [12]. Similar study conducted by Divyashanthi and Manivannan revealed, [13] psoriasis and contact dermatitis were most common dermatological disorder in which corticosteroid prescribed.

In this study, most common corticosteroids prescribed were betamethasone 19%, Clobetasol proprionate 17%, and Deflazacort 15%. Similar findings revealed in Javsen et al study [14]. In contrast, a study by Bylappa et al found clobetasol to be the most widely utilized corticosteroid. Similar to this study, Haiya J S. et al found that fluorinated glucocorticoids Betamethasone and Clobetasol were the most regularly prescribed, whereas non-fluorinated glucocorticoids Prednisolone and Mometasone were the least usually administered [15]. Fluorinated steroids have a higher permeability and produce systemic adverse effects due to their higher lipid solubility. In this study, corticosteroid mainly prescribed alone (85%). Most commonly used topical steroid combination with Fusidic acid 4%, Gentamycin 1%, and Miconazole nitrate 5% and with Clotrimazole 5%. Similarly to the current study, Nerurkar RP et al found that the 88% of prescribed corticosteroids were topical as single drugs, while only 12% were recommend in combinations with other topical medicine such as Salicylic acid, Fusidic acid and Clotrimazole [16]. Various studies conducted in various nations have also found comparable results to the current study [12, 17]. Only if the lesion was limited to a tiny area of skin were topical steroid fixed dose combinations are preferred. A short course of an appropriate oral anti-infective was chosen in severe instances [15].

As a result, the topical route was proven to be the most chosen route for medication administration (62 %) in the current investigation. When compared to other studies, one consistent conclusion was that topical formulations were widely utilized in dermatology [10, 18-19]. The principal advantage of adopting the topical route for steroid administration was that it had the fewest side effects, unless systemic injection was required [3, 20]. Unless the disease requires long-term treatment, oral formulations should be administered in a short period with reducing doses [21].

According to the results of the current study, nearly 51% of the steroids administered were very potent, while just 26% were low potent. In terms of duration of use, 67 % had been on corticosteroids for more than two weeks. According to a study by Bhagunde et al, high potency medications were prescribed in 66% of encounters, whereas low/mild potency corticosteroids were prescribed in 11% of encounters [7]. Other studies have shown similar results [15,22]. Long-term and extreme application may result in hypothalamus pituitary adrenal axis suppression as well as local side effects [22,23]. When at all possible, the use of highly potent corticosteroids should be restricted.

Low potency corticosteroids have been considered to be the best for long-term use [24, 25]. High potency corticosteroids have been recommended over low and moderately potent corticosteroids in specific situations, such as atopic dermatitis [26].

This study had few drawbacks. Patients were not followed up on in this cross-sectional trial to assess the efficacy and safety of corticosteroids. Only dermatological out-patient department were included in the study. Inpatients and emergency situations all required to be investigated. Future studies will be able to assess this. Maintaining a balance between judicious use and frequent misuse of corticosteroids, as well as physician attention and patient education during consultation is crucial.

## Conclusion

More emphasis should be placed on rational and comprehensive corticosteroids prescribing for skin diseases. The baseline data generated by these studies can be used by researchers and policymakers to enhance prescribing practices. Continuing medical education is also critical for professional physicians to put it into action and ensure success. This will help to improve corticosteroid prescribing by providing education during consultations.

## Data Availability

yes

